# Antigen-based rapid diagnostic testing or alternatives for diagnosis of symptomatic COVID-19: A simulation-based net benefit analysis

**DOI:** 10.1101/2020.12.16.20248357

**Authors:** Emily A Kendall, Nimalan Arinaminpathy, Jilian A Sacks, Yukari C Manabe, Sabine Dittrich, Samuel G Schumacher, David W Dowdy

**Author notes:** Co-senior authors contributed equally (Samuel G Schumacher, David W Dowdy).

## Abstract

**Background:** SARS-CoV-2 antigen-detection rapid diagnostic tests (Ag-RDTs) can diagnose COVID-19 rapidly and at low cost, but their lower sensitivity than nucleic acid amplification testing (NAAT) has limited clinical adoption.

**Methods:** We compared Ag-RDT, NAAT, and clinical judgment alone for diagnosing symptomatic COVID-19. We considered an outpatient setting (10% COVID-19 prevalence among the patients tested, 3-day NAAT turnaround) and a hospital setting (40% prevalence, 24-hour NAAT turnaround). We simulated transmission from cases and contacts and relationships between time, viral burden, transmission, and case detection. We compared diagnostic approaches using a measure of net benefit that incorporated both clinical and public health benefits and harms of intervention.

**Results:** In the outpatient setting, we estimated that using Ag-RDT instead of NAAT to test 200 individuals could have a net benefit equivalent to preventing all symptomatic transmission from one person with COVID-19 (one “transmission-equivalent”). In the hospital setting, net benefit analysis favored NAAT, and testing 25 patients with NAAT instead of Ag-RDT achieved one “transmission-equivalent” of incremental benefit. In both settings, Ag-RDT was preferred to NAAT if NAAT turnaround time exceeded two days. Both Ag-RDT and NAAT provided greater net benefit than management based on clinical judgment alone, unless intervention carried minimal harm and was provided equally regardless of diagnostic approach.

**Conclusions:** For diagnosis of symptomatic COVID-19, the speed of diagnosis with Ag-RDT is likely to outweigh its lower accuracy compared to NAAT wherever NAAT turnaround times are two days or longer. This advantage may be even greater if Ag-RDTs are also less expensive.

## Introduction

Accurate diagnosis of COVID-19 can guide clinical management; reduce transmission; and inform appropriate allocation of resources for isolation, contact tracing, and treatment. In many settings – including low- and middle-income countries (LMICs) as well as high-income countries with large outbreaks – efforts to diagnose COVID-19 using nucleic acid amplification tests (NAATs) frequently exceed capacity.^1–3^

Antigen-detection rapid diagnostic tests (Ag-RDTs) are less expensive than NAATs and can be performed in minutes without centralized laboratory infrastructure. Thus, they could facilitate higher volumes of testing and provide rapid results while relieving strains on laboratory capacity. Ag-RDTs are, however, less sensitive than NAATs. Some experts have argued that all SARS-CoV-2 testing must be highly sensitive, ^4,5^ while others advocate less sensitive testing primarily for community-level surveillance.^6^ Recognizing the limited capacity for NAAT in many settings, WHO and national public health agencies have issued target product profiles for Ag-RDTs^7^ and interim guidance for their use in select circumstances.^8–10^ Meanwhile, a global partnership has begun to manufacture and distribute 120 million Ag-RDTs in LMICs.^11^

As Ag-RDTs become more broadly available, it is important to understand the conditions under which their implementation would be preferable to relying on NAAT and/or clinician judgment alone. This understanding must balance accuracy (i.e., sensitivity and specificity) with other considerations such as the turnaround time for results and the likely actions in response to a positive (or negative) diagnosis. We sought to aid decision-making by quantifying these tradeoffs into a single unified measure of net benefit for patients presenting with symptomatic COVID-19 in the inpatient and outpatient settings.

## Methods

### Overview

We developed a simulation model of COVID-19 diagnostic evaluation, in which we compared the use of point-of-care Ag-RDT, centralized (e.g., hospital-based) NAAT, and clinical judgment (i.e., without virological testing) among individuals presenting with symptoms suggestive of COVID-19. We parameterized the model using published data (and assumptions, where published data did not exist) regarding SARS-CoV-2 viral dynamics, clinical features of COVID-19, and diagnostic assay performance. The model accounts for the following aspects of disease dynamics and diagnostic testing: (1) time-dependent transmission from index cases and their contacts (Figure 1A); (2) variable timing of clinical presentation; (3) non-uniform distribution of peak viral burden across the population (Figure 1B); (4) correlation of viral burden with both assay sensitivity (Figure 1B, dotted horizontal lines) and infectivity (Appendix Figure A1); and (5) assay-dependent decline in sensitivity with time since symptom onset (Figure 1C). After simulating these dynamics and their effects on transmission-related and clinical outcomes, we used an adaptation of net benefit analysis^12–14^ to combine the transmission-related benefits, the clinical benefits, and the harms of intervention after diagnosis on a single net-benefit scale with units of “transmission-equivalents,” where 1.0 = equivalent benefit to averting all symptomatic transmission from an average person with symptomatic COVID-19. We then used this scale to characterize the incremental net benefit of Ag-RDT relative to either NAAT or clinical judgment alone, under a variety of assumptions. While costs can also be included in calculation of net benefit, this requires monetization of clinical and transmission benefits; thus, we elected to exclude costs from this analysis.

**Figure 1:**
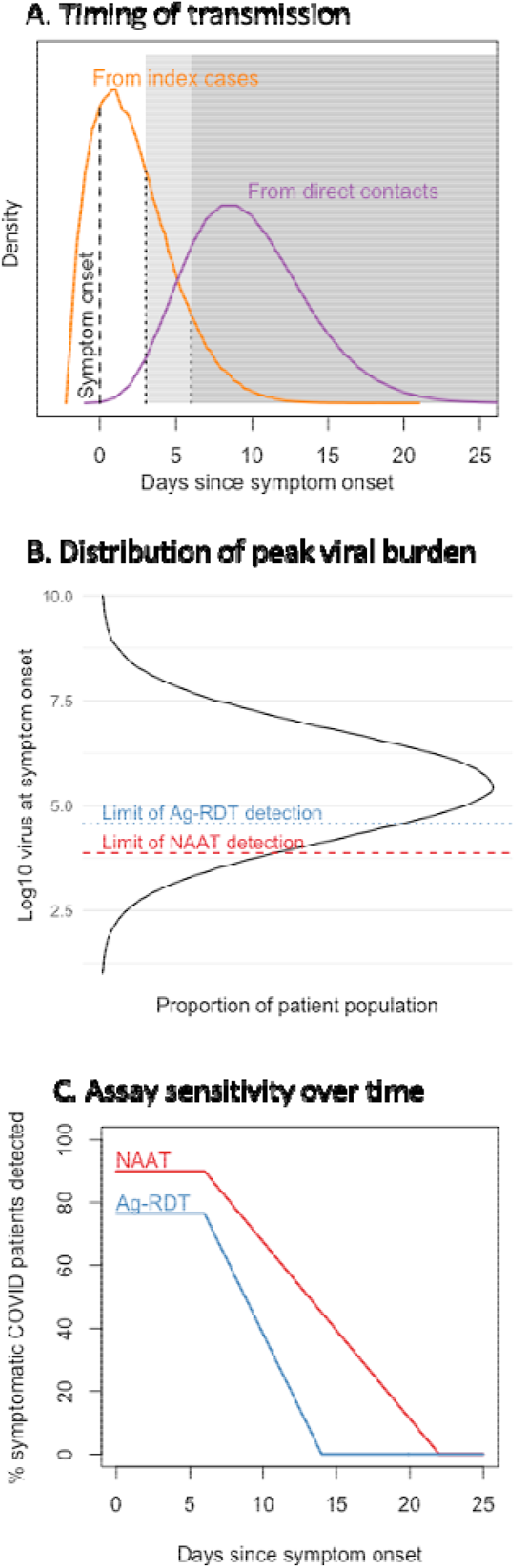
Key assumptions in simulation of net benefit. **Panel A**: The timing of transmission events both from cases and from their direct contacts, in absence of intervention, follows a non-uniform distribution relative to the time of symptom onset in the index case. Shading illustrates that different amounts of transmission are preventable after earlier (light shading) versus later (dark shading) diagnosis. **Panel B**: The simulated viral burden in upper respiratory diagnostic specimens at symptom onset is log-normally distributed across a simulated population. This may be conceptualized in terms of genome copies on a quantitative NAAT (units shown on y axis) or antigen copies (assumed to vary in proportion to genome copies during acute illness). **Panel C**: After an initial period of maximal detection, the simulated sensitivity of Ag-RDT declines on a similar timeline as infectivity, and sensitivity of NAAT declines more slowly.

### Simulated patient populations and testing strategies

We separately simulated two symptomatic patient populations being evaluated for COVID-19: patients with moderate-to-severe illness in a hospital setting (of whom 40% had COVID-19), and mildly symptomatic people in an outpatient setting (with more nonspecific symptoms, of which 10% represented true COVID-19^15^). As described in the Appendix, other differences between the two settings included a longer turnaround time for NAAT in the outpatient setting (1 day in the hospital, 3 days for outpatients^16,17^), a greater amount of presumptive isolation while awaiting diagnostic results in the hospital setting, a longer mean duration of symptoms before presentation in the hospital setting, and a greater value placed on clinical improvement in the hospital setting versus reduction of transmission in the outpatient setting. These features were all evaluated in sensitivity analyses.

We also modeled differences between NAAT and Ag-RDT assays, based on available data from published studies and preprints. Ag-RDT was assumed to have lower sensitivity at symptom onset (85% relative to NAAT, Appendix Table A1) but faster turnaround time (3 hours, versus 1-3 days for NAAT). In addition to comparing Ag-RDT to NAAT, we compared both virological assays to a clinical (i.e., non-virological) diagnostic approach. Clinician judgment was estimated to have 80% sensitivity (comparable, at the time of clinical presentation, to the proportion of patients detectable by NAAT), with corresponding 50% specificity^18^ (compared to 99.5% and 99%, respectively, for NAAT and Ag-RDT, Appendix Table A1). We assumed that clinical diagnoses, being less certain, may result in less intense intervention – for example, less stringent isolation, less case reporting or contact notification, or more circumspect clinical management – and therefore reduced both the benefit and harm of intervention (by a factor of 0.75 in the primary analysis), compared to virological diagnoses made using Ag-RDT or NAAT. For directness of comparison, we assumed that only a single diagnostic approach would be used in a given setting and patient population, with no further testing or clinical risk stratification after the initial evaluation.

### Simulated SARS-CoV-2 transmission dynamics and diagnosis

Our model stochastically simulated the timing of transmission events from index cases and their contacts, relative to index cases’ symptom onset (Figure 1a; details in Appendix). The model accounted for pre-symptomatic transmission,^19–21^ delays between symptom onset and clinical presentation,^22–24^ partial isolation while awaiting test results, and more stringent (but still imperfect) isolation and contact tracing/quarantine once a COVID-19 diagnosis was made.^25,26^

We assumed that viral burden coincided with the onset of symptoms and was log-normally distributed in the population (Figure 1b),^27,28^ and that both diagnostic sensitivity and infectivity were functions of this viral burden. Specifically, based on observed relationships between NAAT cycle threshold, viral culture, and infectivity,^22,29–31^ and similar to other models,^6^ we assumed a log-linear relationship between peak viral burden and peak infectivity, above a minimum threshold of 10^3^ viral genome copies. We also assumed that for both Ag-RDT and NAAT, infected patients who tested positive had higher viral burdens than those who tested negative (Figure 1b). Thus, infectivity was greater for NAAT-positive/Ag-RDT-positive patients than for NAAT-positive/Ag-RDT-negatives (Appendix Figure A1). We also assumed that assay sensitivity declined over time after an initial six-day period of maximal sensitivity; Ag-RDT sensitivity was assumed to decline on a similar timeline as infectivity, while NAAT sensitivity declined more slowly and thus detected some patients who were no longer infectious (Figure 1c; details in Appendix).^22,32–34^

Using these assumptions, we simulated a population of “index” patients with COVID-19, each with a specified peak viral burden and time to clinical presentation. We chose to simulate a large population of 1,000,000 index patients, to reduce stochastic variability and provide more reliable estimates of mean behavior (Appendix Table A4). (Additional patients without COVID-19 enter our analysis at a later step when we estimate the harms of false-positive results.) We simulated the timing of “first-generation” transmission events from index patients to their direct contacts (according to index patients’ relative infectivity) and of “second-generation” transmission events from index patients’ contacts to contacts of their own, in absence of intervention. We then simulated the result of each diagnostic test, based on each patient’s peak viral burden, the time from symptom onset to testing, and assay sensitivity at that time (Figure 1C). Finally, we simulated the effects of case isolation and contact quarantine as probabilities of averting first- and second-generation transmission events, respectively (Appendix Table A2).

In addition to the transmission impact of isolation and quarantine, we also modeled the clinical impact of COVID-19-specific medical treatments as time-sensitive. Specifically, we assumed that earlier treatment had greater impact on morbidity and mortality, declining exponentially with time since symptom onset. We also assumed that the average impact of treatment was much greater in the hospital than the outpatient setting (Appendix section 2.1).

### Estimating benefit of diagnostic approaches

We created a unified measure of the combined value (“net benefit”) of each testing strategy, equal to (a) the public health benefit of preventing transmission through case isolation and contact quarantine, plus (b) the clinical benefit of preventing morbidity and mortality through COVID-19-specific treatment, minus (c) the harms of these interventions. To combine these quantities on a common scale, we placed these benefits and harms on a scale of “transmission-equivalents”, where a value of 1.0 equals the value of preventing all symptomatic transmission from one patient of average infectiousness who develops symptoms. Our novel approach builds on decision curve analysis, in which benefits and harms of intervention are estimated on a single scale using a weighting factor. ^14,34,35^ Two novel ways in which our approach builds on this framework are (i) separately estimate clinical and public health benefits in a mechanistic way, and (ii) capturing how benefit may vary depending on the patient or the timing of diagnosis. In doing so, our analysis therefore captures key trade-offs between the accuracy and timeliness of diagnosis, trade-offs that are inherent in comparing Ag-RDTs with NAAT. Our analysis also differs from the conventional approach in that it incorporates potential harms of intervention (e.g. inconvenience of isolation, side effects of treatment) for both true-positive and false-positive diagnoses.

Transmission-related benefits were estimated as a weighted average of the proportion of transmission that was prevented from index cases (through isolation) and from contacts (either by preventing infection or through contact quarantine) (Appendix section 1.6). This quantity was estimated relative to the total amount of transmission that was modeled to occur after symptom onset, to estimate benefit in “transmission-equivalents.” Clinical benefits were estimated by first estimating the effectiveness of a given diagnostic approach at averting avertible mortality, accounting for delays in diagnosis and treatment and for imperfect test sensitivity (Appendix Section 2.1). This benefit was converted to transmission-equivalent units by estimating the number of cases from whom all symptomatic transmission (to average-risk individuals) would need to be prevented, in order to achieve the same expected mortality reduction as would be achieved through prompt treatment of one symptomatic case in the setting under consideration (Appendix section 2.1).

The estimated per-patient transmission-related and clinical benefits were multiplied by the prevalence of COVID in the population (i.e., assuming no benefit to patients who had symptoms but no underlying COVID-19), to determine the benefits of testing per patient tested. These benefits were then reduced by an estimate of the per-patient harms of testing and resulting intervention (e.g., inconvenience of isolation or side effects of treatment). The harms were assumed to apply to every patient with a positive diagnostic result regardless of whether that patient had underlying COVID-19. To place harms on the same transmission-equivalent scale as benefits, we used an approach from decision curve analysis:^35^ we defined a “threshold probability” *q* of a patient having a condition (in this case, COVID-19), above which one would isolate a newly symptomatic patient and expect the benefits to outweigh harms. In our formulation, this corresponds to an assumption that the benefit of intervening promptly on a patient with COVID-19 is 1/*q* times as large as the harm of intervening on any patient (irrespective of COVID-19 status); for purposes of our analysis, therefore, we refer to this quantity *q* as the “harm-benefit ratio”. We estimated *q* at 0.1 in the reference scenario based on the observed willingness to isolate or quarantine individuals in current practice (Appendix section 2.2); but as is standard in decision curve analysis, we also considered results across a range of harm-benefit ratios. The resulting net benefit equation is presented and explained in Appendix section 2.3.

We performed both one- and two-way sensitivity analyses of our model parameters, varied across the ranges specified in Appendix Table A1. In these analyses, we estimated the net benefit of each diagnostic approach and the incremental net benefit of Ag-RDT relative to NAAT. We also present decision curves that show the net benefit of each strategy according to the harm-benefit ratio *q*.

This research did not involve human subjects and did not require ethical review. All analyses were performed using R version 4.0.2.

## Results

### Transmission-related benefits of diagnosis

The simulated number and timing of transmission events, under each diagnostic scenario, are shown in Figure 2. In the outpatient setting, an estimated 63% of COVID-19 cases were detected under the Ag-RDT scenario, compared to 80% for both NAAT and clinical judgment (Figure 2A); in the hospital setting, later presentation slightly reduced these percentages, to 57% and 75%, respectively (Figure 2B). However, Ag-RDT detected cases more rapidly than NAAT (Figure 2A-B), while also detecting the cases who were most infectious (for example, the 63% of patients who were Ag-RDT-detectable accounted for 77% of all subsequent transmission events). These effects led to prevention of similar amounts of transmission by Ag-RDT (blue line, Figure 2C-D), NAAT (red line, Figure 2C-D) and clinical judgment alone (with less intensive intervention; green line, Figure 2C-D). In the hospital setting, NAAT remained the most effective approach for averting transmission (Figure 2D), but in the outpatient setting (Figure 2C), we estimated that Ag-RDT could avert 32% of all transmission, compared to 29% for NAAT and 31% for clinical judgment alone.

**Figure 2:**
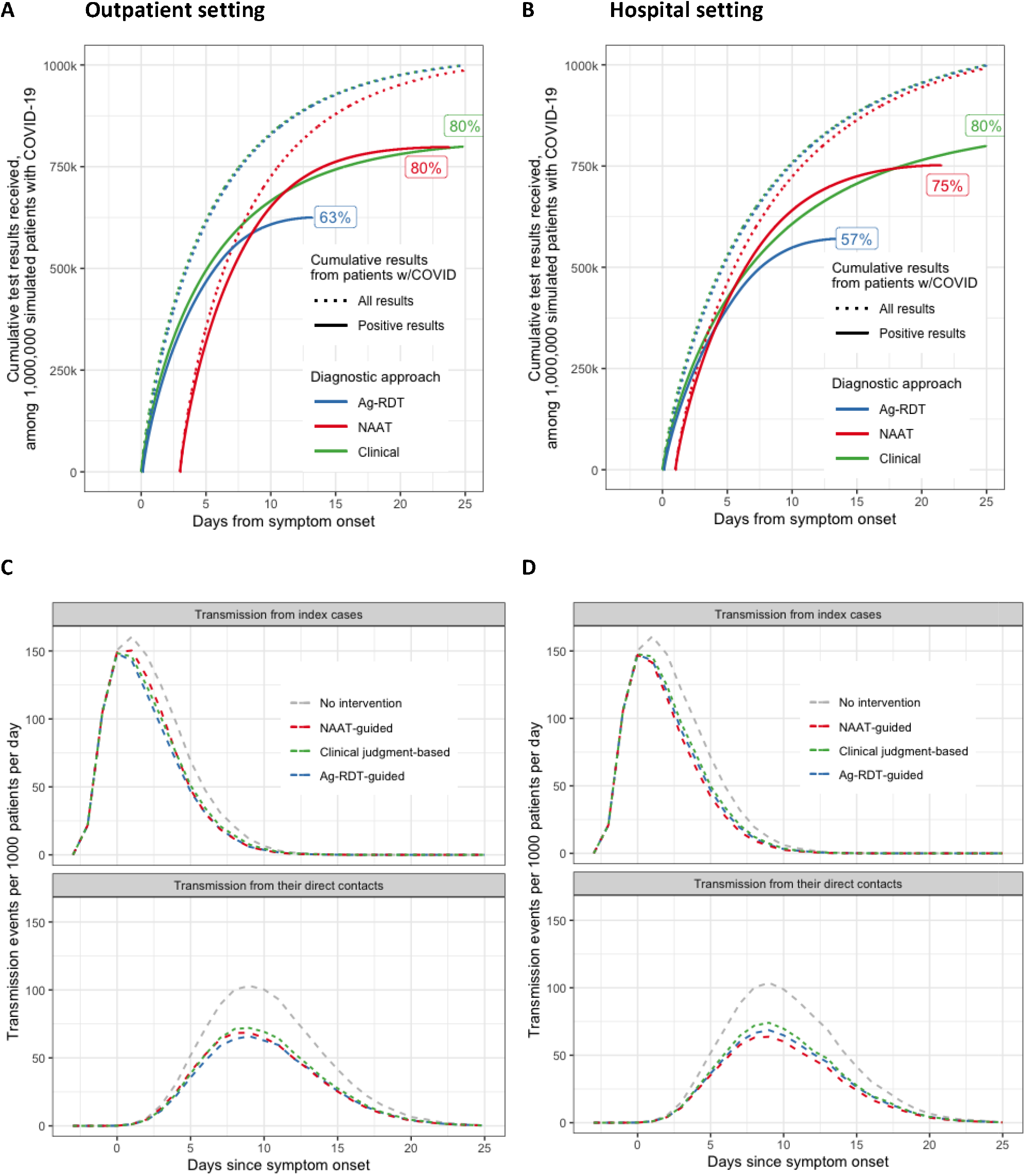
Timing of testing and SARS-CoV-2 transmission events under different diagnostic algorithms in the outpatient setting. **Panels A-B:** Cumulative test results over time among 1,000,000 patients with COVID-19 in the (A) outpatient and (B) hospital settings. Time of symptom onset and clinical presentation (3 days after symptom onset) is aligned for all patients; thus, NAAT turnaround time is depicted as the space on the x-axis before the start of the red curves. Solid lines indicate all results received (including false-negative results) from patients with COVID, while dashed lines indicate only the positive results. Ag-RDT and clinical judgment provide same-day results (and thus have curves shifted to the left) relative to NAAT, but NAAT ultimately detects more true-positive COVID-19 cases than Ag-RDT (80% versus 63%). Clinical judgment may diagnose as many true COVID-19 cases as NAAT, but with lower specificity and no preference for the most infectious cases (not shown). **Panels C-D:** Quantity and timing of transmission with and without intervention, in the (C) outpatient and (D) hospital settings. Delayed clinical presentation, incomplete isolation, and incomplete contact notification/quarantine prevent any testing strategy from preventing the majority of transmission (difference between grey lines and colored lines). Ag-RDT (blue lines) achieves slightly greater reductions in transmission in the outpatient setting where NAAT delays are long, whereas NAAT (red lines) achieves slightly greater reduction in the hospital, where the benefit of greater sensitivity outweighs the negative effect of a one-day delay in results.

### Net benefit, incorporating harms and clinical benefits of diagnosis

Figure 3 shows estimates of net benefit for Ag-RDT relative to NAAT, under different scenarios of NAAT turnaround time and Ag-RDT sensitivity (relative to NAAT). In the outpatient setting, the net benefit provided by either NAAT or Ag-RDT testing, relative to no testing, was estimated to be between 0.02 and 0.04 transmission-equivalents (Figure 3A-B, lower dotted lines). Thus, 25 to 50 individuals would need to be tested to achieve a net benefit equivalent to preventing all symptomatic transmission from one average case with symptoms. In the hospital setting, where the prevalence of COVID-19 was higher and the potential benefits of prompt treatment much greater, the net benefit of testing (relative to no testing) ranged from 0.3 to 0.6 transmission-equivalents (figure 3A-B, upper solid lines). Thus, between two and three tests in the hospital were needed to achieve the same net benefit as 25 to 50 tests in the outpatient setting.

**Figure 3:**
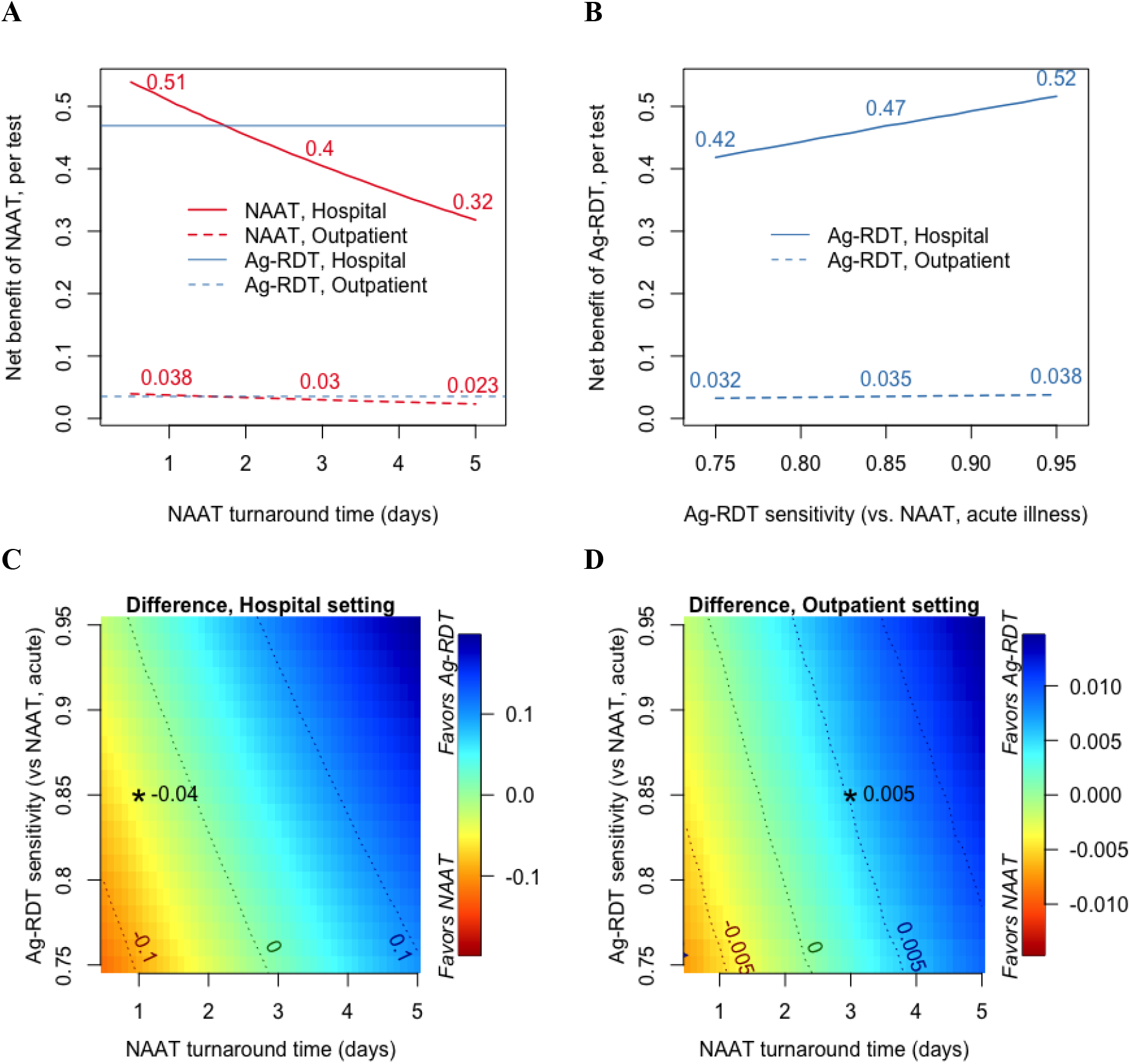
Net benefit of COVID-19 testing strategies. Panels A and B show the net benefit of testing each patient population with NAAT (**panel A, red**) or Ag-RDT (**panel B, blue**) over a range of values for NAAT turnaround time (panel A) or assay sensitivity (panel B). Net benefit is expressed in “transmission-equivalents” per patient tested, with 1 unit equivalent to the value of preventing all symptomatic transmission from one average case. Points of intersection in panel A indicate equivalent net benefit for the two testing strategies, holding all other parameters at their reference values. Panels C and D show the difference in net benefit between the two assays (Ag-RDT minus NAAT) in the outpatient setting (**panel C**) and hospital setting (**panel D**). The colors scales differ by setting, due to the larger magnitude of net benefit per patient tested, for either test, in the hospital setting. Asterisks (*) indicate the net benefit associated with our primary estimates of NAAT turnaround time and Ag-RDT sensitivity for each setting, and contour lines indicate parameter value combinations that produce the same specified incremental net benefit.

In both settings, the points at which Ag-RDT provided incremental net benefit relative to NAAT (i.e., points of intersection in Figure 3A, “0” contour lines in Figures 3C-D) were sensitive to the turnaround time for NAAT results (Figure 3A) and, to a lesser degree, the sensitivity of Ag-RDT (Figure 3B). At our primary estimates of Ag-RDT sensitivity (85%, defined acutely relative to NAAT), Ag-RDT provided an incremental net benefit of 0.005 transmission-equivalents relative to NAAT in the simulated outpatient setting with an assumed three-day NAAT turnaround time (Figure 3C, asterisk), whereas NAAT provided an incremental net benefit of 0.04 transmission-equivalents relative to Ag-RDT in the simulated hospital with an assumed one-day NAAT turnaround time (Figure 3D, asterisk). In both settings, assuming Ag-RDT sensitivity of 85% relative to NAAT, Ag-RDT was the preferred strategy (i.e., positive incremental net benefit) when NAAT turnaround times were ≥2 days. This threshold for equivalency dropped to one day if Ag-RDT was assumed to have 95% sensitivity, and increased to three days if Ag-RDT sensitivity was assumed to be 75% relative to NAAT (Figure 3C-D, “0” contour lines).

Our finding that Ag-RDT offered positive incremental net benefit relative to NAAT in the outpatient setting was robust to variation in most other parameter values (Figure 4A). One exception was isolation practices while awaiting test results: if strict isolation (i.e., sufficient to reduce transmission by 70%) were maintained while waiting for NAAT results, without any additional harms, then NAAT would be preferred to Ag-RDT in this setting. In the hospital setting (Figure 4B), Ag-RDT could offer greater net benefit than NAAT if clinical interventions were highly time-sensitive (such that the morbidity and mortality that remained avertable by treatment decreased by 30% per day). Contributions of prevented transmission, prevented mortality, and false-positive diagnoses to the net benefit are explored in Appendix Table A3.

**Figure 4.**
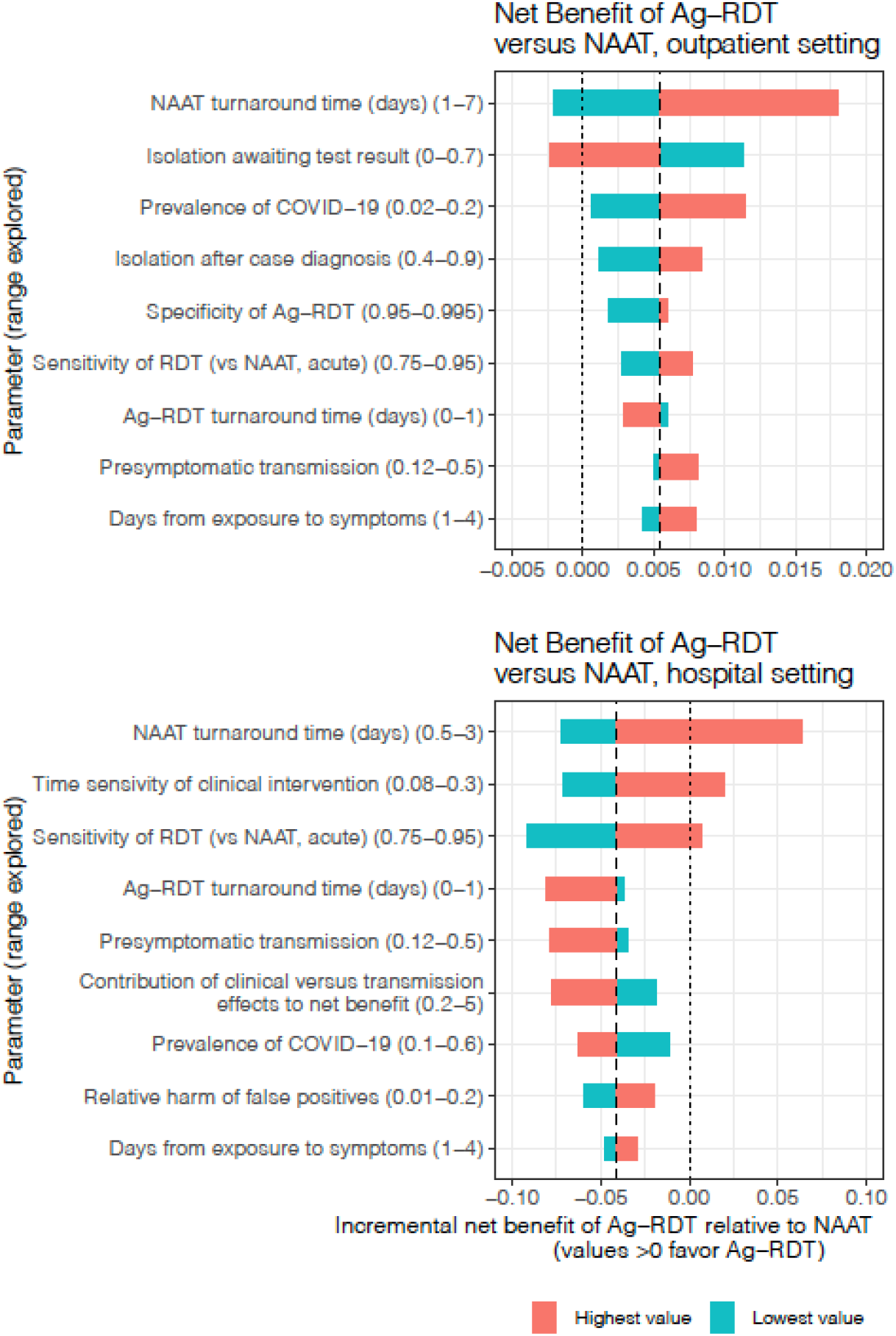
One-way sensitivity analysis for incremental net benefit of Ag-RDT compared to NAAT, in (A) the outpatient setting and (B) the hospital setting. The incremental net benefit associated with a given parameter value, holding all other parameters fixed at their primary estimates, is compared to the primary estimate of incremental net benefit, in units of transmission-equivalents per patient tested. Dotted vertical lines at x=0 mark the point of equivalent net benefit between Ag-RDT and NAAT, such that bars which cross this line indicate a change in the conclusion about which test offers greater net benefit. All parameters were explored; only those associated with >25% change in the estimate of incremental net benefit (for the high and/or low value of the parameter) are displayed.

As a final consideration, the estimated net benefit of each assay was estimated to change as the harm of intervening on a positive result, relative to the benefit of prompt intervention in a patient with COVID-19 (i.e., “threshold probability” for treatment) increases. Red and blue lines in Figure 5 illustrate that, at high relative harms of intervention, Ag-RDT is increasingly preferred over NAAT, because the harms of intervening on NAAT-positive/Ag-RDT-negative individuals (who are less infectious than average or late in their disease course) become large relative to the benefits. By contrast, at very small estimates of relative harm, clinical judgment – with high sensitivity but low specificity and thus high probability of false-positive diagnosis – becomes increasingly favored. However, the net benefit of clinical judgment was projected to exceed that of NAAT or Ag-RDT only if clinically diagnosed patients could receive interventions of equal intensity as patients with a positive virological test (dotted versus solid green lines in Figure 5) – and even then, only if the harm of such intensive intervention was deemed equivalent to 10% or less of the benefit of intervening on one person with true COVID-19.

**Figure 5.**
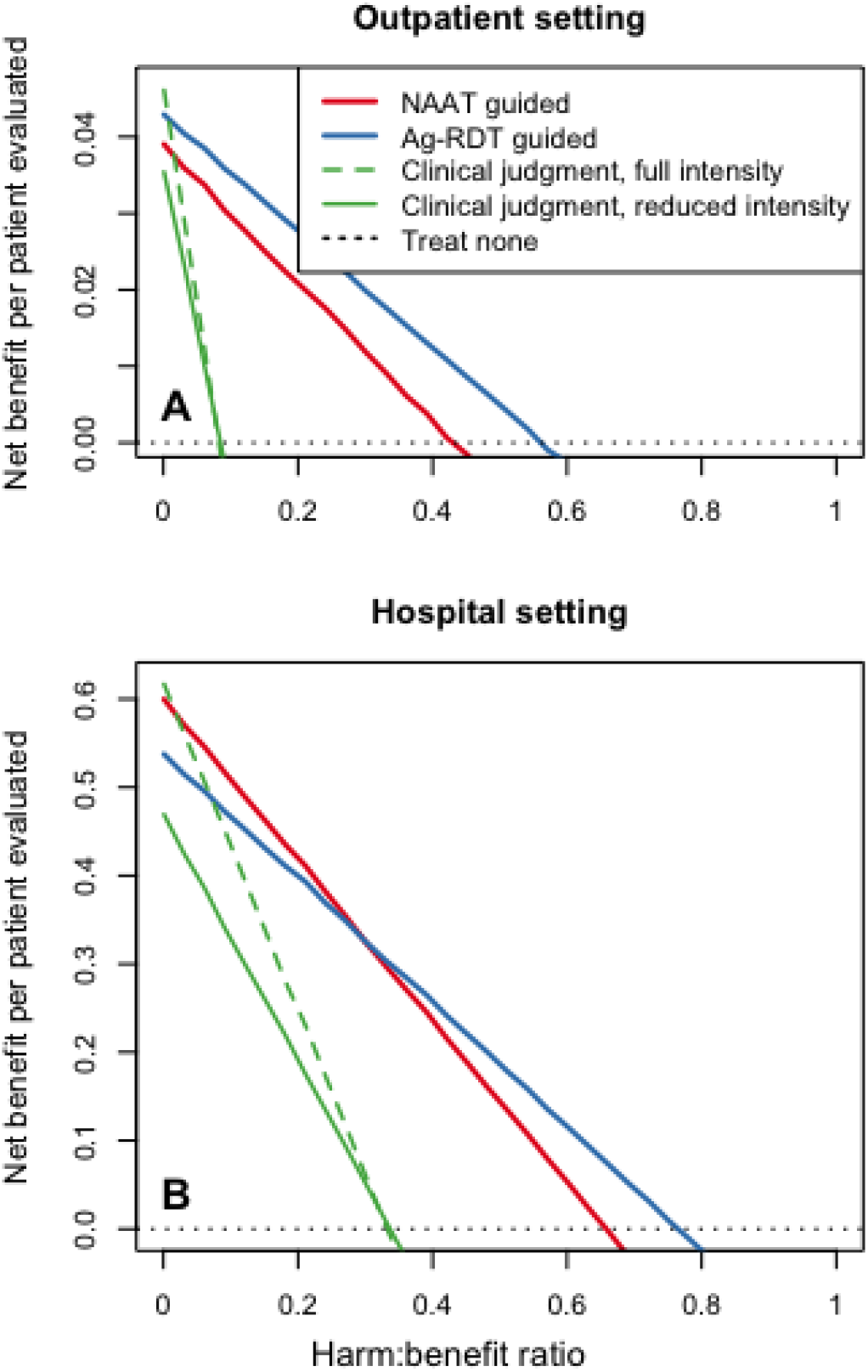
Comparing NAAT, Ag-RDT, and a clinical approach over a range of estimates for the harms of intervention relative to its potential benefit. In the clinical approach (green lines), intervention is guided by clinical judgment alone, but may be provided at a less-effective intensity (dashed green lines) due to greater uncertainty. The x-axis depicts the ratio of the harm associated with intervening on a positive test result (regardless of underlying COVD-19 status) to the benefit of intervening promptly (i.e., at symptom onset) in someone with COVID-19. The same harm is assumed to apply to all interventions (in contrast to a traditional decision curve analysis, which considers harms only to accrue to those with false-positive test results), and thus, the harm:benefit ratio is equal to the threshold probability of disease at which the expected harms and benefits of intervention are balanced. Net benefit is estimated in “transmission equivalents” as defined above. If clinical diagnoses receive the same full intervention as virologically diagnosed cases (dashed green lines), then clinical judgment could outperform Ag-RDT and NAAT if the relative harm of intervention is very low. If, however, the lower certainty of a clinical diagnosis results in a reduced intensity of intervention and therefore reduced benefit (by 25% in our primary analysis, solid green lines), then virologic testing is likely to provide greater net benefit even if the harms of intervention are negligible.

## Discussion

As antigen-detection SARS-CoV-2 rapid diagnostic tests become more widely available, it is important to identify the settings under which they offer incremental benefit over NAAT or clinical judgment alone. We developed a novel adaptation of net benefit analysis that accounts for both the clinical and transmission-related benefits of diagnosis in clinical settings, as well as the harms of intervening (on both false-positives and true-positives). Using this framework, we demonstrate that Ag-RDT is likely to yield greater health benefit than NAAT under typical outpatient conditions that include turnaround times of ≥2 days for NAAT results, even if Ag-RDTs have considerably lower sensitivity. We demonstrate that Ag-RDT would be similarly favored over NAAT in a hospital setting with NAAT turnaround-times of 2 days or more – though faster turnaround times are likely to be available in many hospitals. Furthermore, an Ag-RDT assay that achieved sensitivity >95% relative to NAAT during acute illness could provide greater benefit than NAAT even at NAAT turnaround times of one day. Finally, we also demonstrate that both NAAT-based and Ag-RDT-based testing are preferable over clinical judgment alone, under reasonable assumptions about the harms of false-positive diagnoses and the likely intensity of response to virologically unconfirmed diagnoses.

Compared to other estimates of the transmission burden that might be prevented through frequent asymptomatic screening^37–39^, our results suggest that symptom-driven testing may have limited potential to reduce transmission, because of late clinical presentation relative to the onset of infectiousness. (One exception, not modeled here, might be settings with extensive backward contact tracing after a symptom-based diagnosis ^40,41^). Our results also show that testing alone will avert only a small proportion of poor clinical outcomes, due to the limited efficacy of COVID-specific treatments and the frequent empiric provision of nonspecific treatments. Nevertheless, our comparisons of NAAT and Ag-RDT in clinical settings align with models comparing the same tests for asymptomatic screening.^6,38^ Specifically, our results indicate that rapid results can overcome the disadvantages of suboptimal sensitivity and allow the net benefit of Ag-RDTs to match or (when NAAT turnaround times are large) exceed that of slower but more sensitive NAATs.

Our novel application of net benefit and decision curve analysis to COVID-19 demonstrates the importance of accounting for factors beyond sensitivity and specificity – including the ability to diagnose the most epidemiologically important and clinically treatable cases – when evaluating infectious disease diagnostics. Incorporating all of these considerations into a single metric of net benefit, our analysis suggests that Ag-RDT is likely to produce better outcomes than NAAT with ≥2 day NAAT turnaround time, as is common in outpatient settings, whereas NAAT is likely to be preferred in facilities with NAAT capacity that can achieve same-day or next-day turnaround. Conventional decision curve analysis – based only on sensitivity, specificity, prevalence, and threshold probability – would incorrectly favor the more accurate test (i.e., NAAT) in all settings, even when the timing of diagnosis is critical.

Although any model-based findings are subject to bias, our estimates are likely to be conservative regarding the benefit of adopting Ag-RDT in clinical settings. First, our analysis does not incorporate the lower economic cost of Ag-RDTs relative to NAAT – though these costs may be partially offset by additional follow-up tests for SARS-CoV-2 or alternative diagnoses that may be required after false-negative Ag-RDT results. Second, our primary estimates of Ag-RDT sensitivity in the acute phase (i.e., 85% relative to NAAT) may be conservative.^22^ Third, our model does not account for the potential that patients who test falsely negative by Ag-RDT may still receive some degree of isolation or clinical intervention on the basis of high clinical suspicion.

Our model is limited by underlying data availability about SARS-CoV-2 dynamics. In particular, data on the relationship between viral burden and infectivity remain sparse. Our ability to draw conclusions that apply across settings and assays is also limited by varying clinical and public health practice – for instance, the extent to which symptomatic people self-isolate or contact tracing is performed are likely to vary widely across settings. Estimates of threshold probability are also context specific, depending, for example, on the individual and societal economic costs of interventions such as contact investigation and quarantine in each particular setting. Third, certain parameter estimates – such as the importance of preventing downstream transmission relative to preventing poor clinical outcomes in people currently infected – depend on prevailing epidemic trends more broadly. Preventing transmission may be more important in settings with emerging or widespread transmission and less important in settings with resolving epidemics or imminent widespread vaccination. Finally, our analysis focuses on diagnosis of symptomatic individuals and quarantine of their direct contacts, and does not consider the potential role of Ag-RDT in preventing pre-symptomatic or asymptomatic transmission.

In conclusion, this novel application of net benefit analysis demonstrates that, for individuals with symptoms suggestive of COVID-19, rapid SARS-CoV-2 testing using a rapid Ag-RDT with 85% sensitivity (measured relative to NAAT and during acute symptoms) could offer greater net benefit than either NAAT or clinician-driven diagnosis. We identified a threshold near 2 days for the turnaround time of NAAT results, above which Ag-RDT is expected to offer greater net benefit than NAAT. Use of Ag-RDT may be particularly beneficial in outpatient settings – where testing volumes are often high, NAAT turnaround times are often long, and the benefits of diagnosis are dominated by the potential to reduce onward transmission.

## Data Availability

Primary data used in this analysis is publicly available and cited in the manuscript. Model code is available from the authors upon request.

