## Appendix for "Antigen-based rapid diagnostic testing or alternatives for diagnosis of symptomatic COVID-19: A simulation-based net benefit analysis"

### 1. Model of viral dynamics

#### 1.1 Overview of simulated patient cohort

In brief, we simulated a cohort of $10^6$ patients with symptomatic COVID-19 in a given clinical setting, and $10^6$ contacts who would be infected by those patients in absence of intervention. Characteristics associated with each patient (sampled from distributions specified below in sections 1.1-1.2 and Table A1) included an upper-airway viral burden at symptom onset (which was mapped to a peak infectivity value), a time from symptom onset to clinical presentation, a time of potential transmission to a contact, and a time of onward transmission from that contact. This patient population was resampled with replacement according to the index cases' relative infectivity to create a sample of $10^6$ potential transmission events from index cases and $10^6$ potential transmission events from their contacts. The result of a diagnostic assay depended the assay type, the assay sensitivity at symptom onset, and the time since symptom onset, and the index patient's peak viral burden or infectivity (represented as a quantile) relative to other patients; patients diagnosed by clinician judgment were chosen at random based with a probability equal to the clinical judgment sensitivity. To determine which transmission events occurred, potential transmission events from index cases were sampled with a probability equal to one minus the intensity of isolation in effect at the time of potential event, and potential transmissions events from contacts whose infections had not been prevented were then sampled with probability equal to one minus the intensity of quarantine they were observing at the time of the potential onward transmission event. Intensities of isolation and quarantine depended on the diagnostic result if it had been received, on empiric isolation practices, and on the intensities of isolation and contact tracing in effect for diagnosed cases. Further details are provided in sections 1.2-1.5, and model parameters are provided in Table A1.

#### 1.2 Temporal distribution of transmission events

We randomly sampled the timing of SARS-CoV-2 transmission events from individuals with symptomatic COVID-19 in absence of intervention, relative to the onset of symptoms in those individuals at time t=0. First, using function get.weib.par from the R package rriskDistributions, we fitted a Weibull (chosen for its three degrees of freedom) distribution to have a 0 quantile at time *-sxOnsetDay,* a *presx* quantile at time 0 (such that *presx* was the proportion of transmission occurring before symptom onset), and a 99% quantile at *duration* (Table A1).

We then simulated the timing of one transmission event from each of the $10^6$ contacts infected in the first generation of transmission. We randomly sampled $10^6$ incubation periods from a gamma distribution with a specified mean and standard deviation (Table A1, [1]), and $10^6$ values for the time from disease onset to transmission from the same Weibull distribution used for the index cases (defined relative to a hypothetical day of symptom onset, even for those contacts who never developed symptoms). The timing of transmission from the contact (relative to onset of symptoms in the index case) was then calculated as the sum of timing of transmission from the index case, the sampled incubation period, and the sampled time from the contact's symptom onset to transmission.

**Table A1 Simulation and net benefit model parameters**

| Variable | Description | Estimate (range explored) | Source/Rationale for estimate |
|---|---|---|---|
| $sn_{NAAT}$ | Sensitivity of NAAT for COVID-19 during acute illness | 90% | [2–4]. Declines over time as described in section 1.4. |
| $sn_{Ag\text{-}RDT}$ | Sensitivity of Ag-RDT for COVID-19, relative to NAAT, during acute illness | 85%<br>(75%, 95%) | [5,6]; in acceptable but suboptimal range per WHO TPP [7]; corresponds to absolute sensitivity during acute illness of 76.5%, and declines over time as described in section 1.4.<br>Sensitivity of Ag-RDT is estimated relative to NAAT; all patients who test falsely negative by NAAT are assumed to also test falsely negative by Ag-RDT. |
| $sn_{clin}$ | Sensitivity of clinician judgment for COVID-19 | 80%<br>(70%, 90%) | Estimated based on clinical prediction scores [8], assuming clinicians consider multiple factors and choose a fairly sensitive (and less specific) point along the range of possible diagnostic thresholds. |
| $sp_{NAAT}$ | Specificity of NAAT for current infectious or symptomatic COVID-19 | 99.5%† | Although NAAT is highly specific for SARS-CoV-2 RNA, some patients may have had COVID-19 that recently resolved, with continued shedding of noninfectious SARS-CoV-2 RNA but new symptoms due to another cause. |
| $sp_{Ag\text{-}RDT}$ | Specificity of Ag-RDT | 99% (95%, 99.5%) | [5,9,10] |
| $sp_{clin}$ | Specificity of clinician judgment | 50% (30%, 70%) | Estimated based on clinical prediction scores [8] in conjunction with the sensitivity estimate above, assuming clinicians consider multiple factors and choose a fairly sensitive (and less specific point along the range of possible diagnostic thresholds. |
| $c$ | Maximum clinical benefit achievable through intervention at symptom onset in the patient population under consideration, in units corresponding to the benefit of preventing all transmission (including pre-symptomatic transmission) from one average case. | Hospital:<br>2 [0.2-5.0]<br><br>Outpatient: 0.06 [ 0-0.1] | Hospital scenario: Based on comparison of preventable hospital mortality to overall infection fatality ratio; see section 2.1.<br>Outpatient scenario: Based on lower COVID mortality among all outpatients, combined with higher potential efficacy but less access to outpatient interventions for those eligible (further details in text section 2.1). |
| $q$ | Threshold probability (of true COVID-19) at which you are willing to intervene at symptom onset | 0.10 (0.01-0.20) | Based on estimate of willingness to treat up to 10 individuals as having presumed COVID-19 in order to appropriately isolate or treat one true COVID-19 case – see "*Estimation of threshold probability*", section 2.2. |

| | | | |
|---|---|---|---|
| *d* | Daily decline in the morbidity/mortality avertible by treatment | Hospital: 0.12 (0.08-0.3);<br><br>Outpatient: 0.18 (0.08-0.3) | Exponential decay model; 12% reflects estimated 5-day half life: Among patients with severe COVID, most divergence in clinical trial survival curves happened in first week[11,12], while median time to death of 19 days sets a bound for the latest that meaningful intervention can occur [13]. For outpatients receiving preventive (e.g. monoclonal antibody) therapy, the time window is similarly narrow, based on 3.5-day median time to hospitalization as the window for potential impact. |
| $t_{NAAT}$ | Turnaround time in days, NAAT | Hospital: 1 (0.5 to 3);<br>Outpatient: 3 (1 to 7) | Outpatient characteristic of high-income countries with large outbreaks and attempted widespread NAAT testing [14,15] |
| $t_{AgRDT}$ | Turnaround time in days, Ag-RDT | 0.125 (0-1) | Three hours (conservative; results may be available in <1 hour[7]) |
| $t_{clin}$ | Time to clinical decision | 0 | Assumed |
| sxonsetday | Average days from onset of infectivity to onset of symptoms | 2 (1,4) | [1] |
| presentationday | Days from symptom onset to clinical presentation | Hospital: Median 5 days (3-8)<br><br>Outpatient: Median 3.5 (2-5) | Modeled as a truncated Weibull distribution with shape 0.85 and max 25 days, and scaled based on median.<br>Hospital: [12,16,17] (accounting for time from presentation to randomization in trial data); Outpatient setting: [18,19]. |
| prev | Prevalence of COVID-19 in the population evaluated | Hospital: 40% (10-60%)<br>Outpatient: 10% (2%-20%) | [9] |
| $iso_0$ | Effectiveness of isolation while awaiting a diagnostic result | Hospital 0.7 (0.4-0.9),<br>Outpatient 0.3 (0-0.7) | Outpatient estimates from adherence to isolation when symptomatic combined with imperfect isolation when at home [20,21]. Assumes high adherence to isolation in hospital when COVID suspected. |
| $iso_1$ | Proportion of future transmission from case averted after diagnosis | Hospital 0.9 (0.5-1);<br>Outpatient 0.7 (0.4-0.9) | Assumes better isolation adherence among known cases than among all who are advised to isolate |
| $iso_2$ | Effectiveness of case diagnosis in reducing future transmission from any contacts who are or become infected | 0.4 (0-0.7) | Reflects the intensity and effectiveness of any contact tracing and quarantine that occurs. Estimate assumes only close contacts are notified, quarantine is incomplete, and those who adhere remain unable to avoid all contacts.[20,22] |
| $l_{clin}$ | Proportion of benefit of a true-positive that is attainable with clinical diagnosis | 0.75 (0.5 – 1.0) | Assumed; varied in sensitivity analysis. Floor value is the approximate reduction in post-test probability after a positive result, comparing a highly specific assay to one with likelihood ratio ~2. Varied in sensitivity analysis. |
| incubation | Incubation period, days | Mean 5.5 (3-8) | [23]; Modeled as gamma distributed, sd 1.5 days. |
| presx | Proportion of transmission that is pre-symptomatic, in a person who develops symptoms but is never diagnosed | 0.2 (0.12-0.5) | Distributions in transmission timing with >40% pre-symptomatic have been estimated, but these are for settings with contact tracing and isolation reducing transmission after diagnosis [1,24]. We are estimating transmission that would occur if not diagnosed. Lower bound of 12% [25] |

| | | | |
|---|---|---|---|
| duration | Time from symptom onset to end of infectivity in 99% of cases | 10 days (5-12) | [26–28] |
| rdt_duration | Time from symptom onset to 0% Ag-RDT sensitivity | 14 days (10-18) | [18,29,30] |
| $v_1$, $v_2$ | Inner 95% quantile range of viral burden on day of symptom onset | $10^3$, $10^8$ | [31,32] |
| *i* | Minimum infectious viral burden | $10^3$ ($10^2$ - $10^5$) | Based on relationship between quantitative cycle thresholds and positive viral culture [33,34]. Primary model increases infectivity log-linearly with viral burden above this threshold. |
| iscale | Skew of infectivity, relative to log(viral burden) at symptom onset | 1 (0-3) | Infectivity = $(\log(v_i/i_1)/\log(\max(v)/i_1))^{iscale}$ if $v_i >= i_1$; 0 if $v_i < i_1$.<br>Value of 1 thus corresponds to a log-linear relationship.<br>Relative infectivity remains proportional over time, for two patients with the same duration of symptoms. |
| $g_1$ | Proportion of transmission benefit that comes from preventing first generation of transmission from case to their direct contacts, versus from preventing downstream transmission | 0.2 (0-1) | Primary estimate represents an average of 4 downstream infections per case. This could reflect a reproductive of 1 with either 20% discounting of each successive generation or a time-limit to widespread transmission (e.g. due to vaccination), or a slowly declining epidemic with reproductive number of 0.8 ($\sum_1^\infty 0.8^i = 4$). The sensitivity analysis range reflects one extreme of equal weight given to index and downstream transmission, and another extreme in which all weight is placed on downstream transmission. |

1.3 Population distribution of viral burden

We modeled both the relative infectivity of patients, and the limit of detection of virologic assays (NAAT and Ag-RDT), as functions of the viral burden in a hypothetical upper-airway diagnostic specimen at the peak of infectivity and the elapsed time since symptom onset. The peak viral burden had a log-normal distribution in each patient population [32,35], with inner 95% quantile as specified in Table A1.

We assumed that this peak viral burden was correlated with the maximal infectivity of the same individual, with the two quantities having a log-linear relationship above a minimum threshold (Figure A1, Panel B); such a relationship has been assumed in other models [36] and is supported by the observed relationship between PCR cycle threshold and probability of culture positivity [33]. In sensitivity analysis, we varied this relationship, ranging from uniform infectivity (iscale=0, Table A1) to a more skewed distribution (infectivity ~ $(\log(\text{virus}))^3$.

We also assumed that such that a NAAT with sensitivity *s* during acute illness would detect those COVID-19 patients whose peak viral burdens were in the upper s quantile of the population distribution (Figure A1).

**Figure A1. Modeled relationship between viral burden, assay detection, and infectivity.** Panel A shows the log-normal distribution of viral burden in diagnostic specimens from 100,000 simulated patients at the time of symptom onset, under the parameter assumptions shown in Table A1. Panel B shows the corresponding distribution of infectivity, assuming a log-linear relationship above viral burdens of $10^3$ copies. An assay with a given sensitivity detects the corresponding upper quantile of viral burdens (e.g. NAAT detects the most infectious 90% of acute cases, white + gray). The dotted vertical lines show the mean infectivity of those detected only by NAAT (blue) and of those detected by both NAAT and Ag-RCT (red). In this model (with iscale=1), individuals detectable by both assays at symptom onset were 2.4 +- 0.9 times as infectious as individuals detected only by NAAT and not by Ag-RDT. Over time, the relative infectivity between two cases remains constant but the sensitivity of each assay declines (not shown).

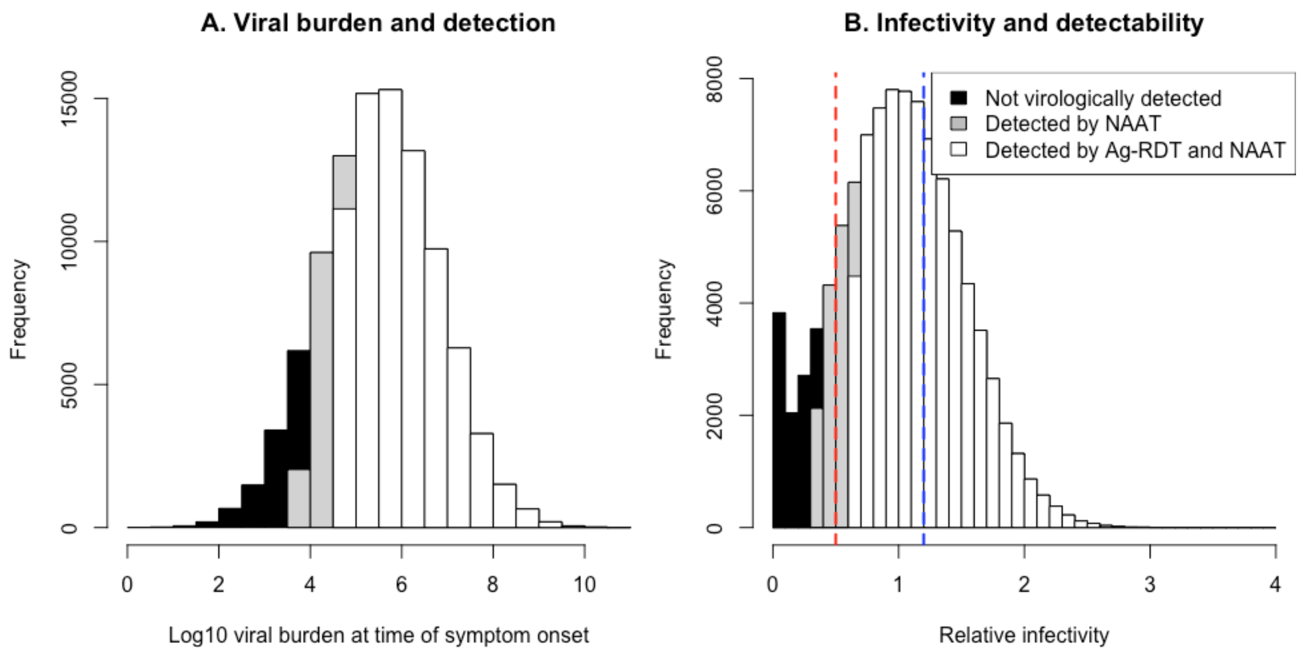

1.4 Virus transmission and detection over time

We simulated the timing of $10^6$ transmission events, in absence of intervention, by sampling the original $10^6$ case transmission events, with replacement, weighted by the relative infectivity of the index cases. Over time since symptom onset, we assumed that relative infectivity between two patients remained constant, such that, for example, if patient A was twice as infectious as patient B on their respective days of symptom onset, then A was also twice as infectious as B on day 5 of their respective illnesses. This, in absence of any intervention, the relative infectivity at the onset of symptoms corresponded to the relative number of secondary cases they generated over the course of their respective illnesses.

For the first 6 days after symptom onset, we assumed that the sensitivity of both NAAT and Ag-RDT remained approximately constant [18,29,37]. After day 6 of symptoms, we modeled sensitivity as declining linearly (based on data from [37] for NAAT, and approximately linear observed relationships between NAAT cycle threshold and time [35,38] and between cycle threshold and probability of a positive Ag-RDT test [39]. We modeled sensitivity as reaching 0 on day 14 for Ag-RDT [18,29] (varied in sensitivity analysis) and reaching 50% of its peak sensitivity value on day 14 for NAAT [37].

1.5 Relationship between diagnostic testing and averted transmission

The timing of diagnosis (which depended on the timing of symptom onset, the time to clinician presentation, and the assay turnaround time) determined which transmission events from index cases or their contacts still might be averted if the case were isolated after diagnosis, or if their contacts were notified and quarantined after diagnosis of the index case. We assumed a high but imperfect degree of case isolation after a positive COVID-19 diagnostic result, and lower intensities of contact quarantine/isolation after case diagnosis and case isolation while awaiting the test result.

We modeled patients as presenting to care and receiving a diagnostic evaluation some number of days after symptom onset, and we modeled each diagnostic approach as having some turn-around time that delayed diagnostic results after the test was performed (Table A1). While awaiting a test result, cases could be partially isolated in a manner that prevented a specified, setting-dependent fraction of transmission events from the index case, but no contact tracing was performed.

Once a diagnostic result was received, those who tested positive were isolated more completely (preventing a setting-specific but high proportion of subsequent transmission events from the index case), and some degree of contact tracing was also implemented (resulting in prevention of a specified proportion of transmission events from contacts of the index case who already were infected or who subsequently became infected).

Thus, the probability that a given diagnostic approach could avert a given transmission event depended on the timing of the transmission event relative to clinical presentation, the turnaround time of the diagnostic assay, whether or not the assay would detect the case (whether the case's viral burden was above the limit of detection for virological assays, and the sensitivity applied as a stochastic probability for clinical judgment), and the intensity of case isolation before and after a result and the intensity of contact tracing and quarantine after a positive result, as shown in Table A2:

**Table A2: Relationship between diagnosis status and prevention of transmission event**

| **Transmission type** | **Timing of transmission event** | **Probability that transmission event is prevented*** |
|---|---|---|
| From case | t < clinical presentation | 0 |
| From case | Clinical presentation < t < diagnostic result | Iso0 |
| From case | t > diagnostic result | If diagnosed, iso1; If not diagnosed, 0 |
| From contact | t < clinical presentation | 0 |
| From contact | Clinical presentation < t < diagnostic result | If transmission from index case to contact was prevented, 1; else, 0. |
| From contact | t > diagnostic result | If transmission from index case to contact was prevented, 1; else, if index case was diagnosed, iso2; else, 0 |

* If diagnosis is made clinically (rather than by NAAT or Ag-RDT), then iso1 and iso2 are reduced by factor $l_{clin}$

1.6 Net Benefit of Averted transmission

For each diagnostic approach, we simulated transmission outcomes and determined what proportion of transmission from cases ($T_{1j}$) (Figure A2), and what proportion of transmission from their contacts ($T_{2j}$), would be prevented by each diagnostic approach *j*. We weighted these two proportions based on an assumed relative value of preventing transmission in the first generation ($g_1$, Table A1); in other words, we assumed that a fraction $g_1$ of the value of prevented transmission arises from avoiding infection of a case's direct contacts, and that fraction 1-$g_1$ is the value of preventing downstream transmission and would be attained even if transmission were interrupted after infection of direct contacts rather than at the index case. Our primary estimate of $g_1$ = 0.2 = 1/5 = $1/(\sum_{i=0}^{\infty} 0.8^i)$ reflects a time-limited epidemic (approaching vaccination, lockdown, or herd immunity within 5 generations of transmission), a discounting of future transmission (by a factor of 0.8), a reproductive number of 0.8 under current epidemiological conditions, or some combination of these assumptions.

We thus estimated the transmission-related benefit, per true-positive diagnosis, of the true-positive diagnoses made with a given assay, relative to the value of preventing all transmission from the average COVID-19 case, as $(g_1)(T_{1j}) + (1 - g_1)\,(T_{2j})$.

Dividing by the proportion of transmission that occurs after symptom onset (*1-presx*, table A1) provides an estimate of the transmission-related benefit of diagnostic testing among patients with COVID, relative to the value of preventing all symptomatic transmission from the average symptomatic COVID-19 case. Multiplying the result by the prevalence of COVID-19 in the patient population, *p*, provides an estimate of the average transmission-related benefit per patient tested, in these same units (i.e., relative to the benefit of preventing all symptomatic transmission from one case.)

### 2. Estimation of weighting parameters and net benefit calculation

#### *2.1 Relative clinical versus transmission benefit*

*Approach:*

We placed the per-patient value of averted poor clinical outcomes on the same net benefit scale as averted transmission – namely, a scale on which one unit corresponds to the value of preventing all symptomatic transmission from one average symptomatic case. We first estimated the number of cases, c, from whom all transmission (including pre-symptomatic transmission) would have to be prevented in order to avert the same amount of eventual morbidity and mortality as could be averted by diagnosing and treating one patient with a given disease severity at symptom onset; for our primary estimates, we estimated c=2 in the hospital and c=0.06 in the outpatient setting, as described later in this section. We divided c by (*1-presx*) to estimate the number of patients whose post-symptom-onset transmission would need to be prevented to achieve this benefit. Finally, we multiplied by the prevalence of COVID in the patient population (*p*), and by the proportion of avertible morbidity and mortality (i.e., avertable at the time of symptom onset) that was averted by a given diagnostic strategy after accounting for the delays from symptom onset to diagnosis $t_{ij}$, and the test result at the time of clinical presentation $x_{ij}$: $\sum_{i=1\ldots n} x_{ij} e^{-d\, t_{ij}} /n$.

*Hospital setting:*

Using US data, we first noted that 6% of diagnosed COVID-19 cases had resulted in hospitalization as of 10/15/2020 [40], assuming 1 week lag. Assuming 5x undercounting on average [41–43], we estimated that this corresponded to hospitalization of 1.2% of all SARS-CoV-2 infections. Among those who are or would be hospitalized, we estimate that prompt COVID-19-directed treatment (care which they would not receive without a COVID-19 diagnosis) starting at the time of symptom onset could avert 10% of their morbidity and mortality, based on observed dexamethasone effect sizes and reductions in in-hospital mortality with improvements in clinical COVID management over the course of the pandemic [12,44].

We then compare this morbidity and mortality avertable through treatment of cases requiring hospitalization, to the morbidity and mortality avertable by preventing all transmission from an average case: Preventing one infection prevents 0.012 hospitalizations and prevents 100% of morbidity and mortality in those hospitalized individuals. Thus, assuming that essentially all COVID morbidity and mortality occur among people who require hospitalization, preventing 8 infections (0.1/0.012) has equivalent morbidity/mortality impact as promptly treating one case who would require hospitalization.

Our estimate of 4 downstream infections per case ($g_1$, Table A1) thus corresponds to c = 2 in the hospital setting. This will be context dependent: in an early and rapidly growing epidemic, preventing all transmission from one average case might ultimately prevent many downstream cases (making *c*<1), while in settings where case numbers are declining or

**Figure A2: Potential net benefit of avertable morbidity and mortality, relative to potential net benefit of preventing transmission**

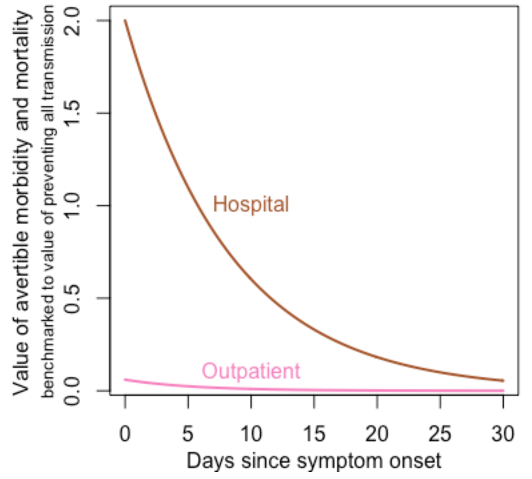

vaccines are being rolled out, contact chains are likely to be saturated or vaccination introduced before 1 case leads to 4 more ($c$>2).

*Outpatient setting*

In the outpatient setting, the estimate of average avertible morbidity and mortality is based on the following assumptions: that most morbidity and mortality occurs in patients who are ultimately hospitalized; that 6% of all diagnosed COVID-19 cases are ultimately hospitalized [40], of whom 50% (3% of all diagnosed cases) are outpatients at the time of initial testing; and that the reduction in morbidity achievable through outpatient management is proportional to the reduction achievable through inpatient management once hospitalized (with the advantages of earlier preemptive therapy, e.g. with treatments that must be given acutely [45] or with more prompt intervention once sick, offset by the lower proportion of at-risk patients who will receive clinical interventions in the outpatient setting even if diagnosed.)

Combining these estimated magnitudes of potential effect with the estimated rates of exponential decay $d$ in avertable morbidity and mortality (Table A1) produces the assumed relationships shown in Figure A2.

### *2.2 Estimation of threshold probability*

We defined the threshold probability for intervention as the probability of COVID-19, i.e., the degree of diagnostic certitude, at which intervening on a patient at the onset of symptoms has zero expected net benefit, because the potential benefit of intervening on a true case is offset by the potential harm of intervening on a patient without COVID-19.

Because we are modeling decision-making for symptomatic patients, and because we are estimating the clinical and transmission benefits from diagnosis in units of prevented post-symptom-onset transmission, we define the threshold probability based on the benefit and risk of intervening at onset of symptoms.

Decision curves plot net benefit across a range of threshold probabilities, but one-way sensitivity analyses around the value of other parameters required us to fix the threshold probability at an estimated value. To do this, we first estimated the threshold probability for the outpatient setting, where the main benefit of diagnosis is ability to interrupt transmission and the main harm of false-positive diagnosis is unnecessary isolation and contact tracing/quarantine. We considered the threshold probability at which, in practice, decision-makers have demonstrated a willingness to isolate or quarantine individuals to prevent SARS-CoV-2 transmission. Such preferences will vary depending on the epidemic growth rate and the social or economic costs of isolation, but as one example, we noted that in local epidemics with largely symptom- or exposure-driven testing and a positive test proportion around 5%, symptomatic people who undergo testing are typically advised to isolate while they await test results [4–6]. Because case diagnosis often results in isolation of more than one individual (case + contacts), we estimated the corresponding threshold for diagnosis of a case at more than 5% probability. On a broader population scale, we noted that in setting with growing epidemics, whole populations have gone on strict lockdown at a prevalence of infectious COVID-19 on the order of 1% (e.g. 100 diagnosed cases/million/day)*(10x undercounting)*(10 infectious days/case) [41,46–48]. Although such lockdown decisions may be based more on projected healthcare resource strain than on prevalence of infectivity, they set a lower bound on the threshold for willingness to isolate populations to prevent transmission. An upper bound is set by household attack rates, where there is consensus that close contacts of a known case (~20% attack rate[49]) should be quarantined. Combining these considerations, we estimated a 10% threshold probability for isolating an individual thought to have COVID in order to prevent transmission from a case of average infectivity, but we note that this estimate is highly context dependent, depending both on the rate of transmission under existing population-wide measures and the additional interventions undertaken for diagnosed cases.

Expanding our estimate to the hospital setting and other clinical contexts where medical treatment might be offered, we judged a similar 10:1 ratio to be a reasonable estimate of the ratio of clinical benefit of appropriate treatment versus clinical harm of false-positive diagnosis (where harms may include side effects of treatments directed to COVID-19 such as steroid or monoclonal antibody treatment, which like the harms of isolation apply to both true- and false-positive diagnoses, as well as delays in other necessary care for wrongly diagnosed patients, e.g. antibiotics for bacterial sepsis). Thus, we inflated the harm per intervention by a factor *1+c* in either setting.

#### *2.3 Calculation of net benefit*

Finally, the estimated transmission-related benefit of diagnosis and resulting public health interventions (section 1.6), the estimated clinical benefit of diagnosis and resulting treatment (section 2.1), and the estimated harm of diagnosis and resulting interventions (section 2.2) were calculated and summed, on a per-patient basis in a patient population with a specific prevalence *p*, and in units corresponding to the benefit of preventing all symptomatic transmission from one case.

The resulting net benefit equation parallels, but is more complex than, the conventional approach. Typically, decision curve analysis calculates net benefit as a weighted difference of the number of true-positive diagnoses (sensitivity * prevalence) and false-positive diagnoses ((1-specificity)*(1-prevalence)); all true-positive diagnoses are assigned equal value, and all false-positive diagnoses are assigned equal harm.[50] Our modified approach similarly combines benefits of intervention for true-positive diagnoses and the harms of intervention for all who receive it, but we use simulation to capture the dependence of those benefits on the patient characteristics and timing of true-positive results, while applying the harm of intervention to both the $p\sum_{i=1\ldots n} x_{ij}$ proportion of simulated patients who test true-positive and the $(1-p)(1-sp_j)$ who have false-positive results, where $sp_j$ is the specificity of diagnostic approach *j*.

Thus, we calculated the net benefit of testing approach *j*, per patient tested, as

$$\left( p\left[\frac{g_1T_{1j} + (1-g_1)T_{2j} + c\sum_{i=1\ldots n}(x_{ij}/n)e^{-d\,t_{ij}}}{(1+presx)} - q(1+c)\sum_{i=1\ldots n}\frac{x_{ij}}{n}\right] - q(1+c)(1-p)\left(1-sp_j\right)\right) l_j$$

Here, $x_{ij}$ indicates whether true case *i* is detected by test *j* at the time of clinical presentation (and thus $\sum_{i=1\ldots n}\frac{x_{ij}}{n}$ represents the sensitivity of test *j* in the population under consideration with *n* simulated true COVID patients); $T_{1j}$ and $T_{2j}$ are the proportions of direct and downstream transmission, respectively, prevented with testing approach *j*; $l_j$ is the reduction in intervention stringency associated with diagnostic uncertainty, which we set to 0.75 for the primary analysis when *j* is non-virological (clinical) diagnosis; and other parameters are as defined in Table A1.

#### *2.4 Estimation of specificity of NAAT*

Even with perfect virologic specificity, in a setting with 50 true infections per 100k population per day, if average 20 day duration of positivity (~10 days beyond end of infectiousness) [51], at least 0.5% of people with unrelated respiratory or viral illness will test “falsely” positive from recently resolved infection.

3. **Estimation of intermediate outcomes:**

To provide context for our net benefit results, we estimate component outcomes of deaths averted (in the patients treated), infections averted (immediate and downstream), and false positives treated, per 1,000 patients tested with each test.

This analysis requires an estimate of mortality that is not part of our original model. As described in Appendix section 2.1, we estimate that prevention of 8 infections has equivalent morbidity and mortality impact as promptly treating one hospitalized case, and that treatment has 30x lower expected impact on mortality in the outpatient setting than in the hospital setting. If we estimate the infection fatality ratio of SARS-CoV-2 infection as 0.5% in the overall general population, then this corresponds to 0.04 deaths averted per patient promptly treated in the hospital setting (i.e., one patient who would die with only supportive care but will be saved by COVID-specific therapy per 25 patients hospitalized with COVID). The mortality prevented by treatment declines with diagnostic delay as specified in our primary model. In the outpatient setting, prompt treatment would avert one death per 750 patients diagnosed.

With these estimates, we obtain the following results for the individual components of the effects of diagnostic testing:

**Table A3: Components of net benefit estimates, each expressed per 1,000 patients tested**

| | **Hospital setting** | | | **Outpatient setting** | | |
|---|---|---|---|---|---|---|
| | **Ag-RDT** | **NAAT** | **Clinical (75% intensity intervention)** | **Ag-RDT** | **NAAT** | **Clinical (75% intensity intervention)** |
| **Deaths averted (in index patients with COVID)** | 15 | 17 | 13 | 0.5 | 0.4 | 0.4 |
| **Infections averted in immediate contacts of index cases** | 182 | 215 | 152 | 178 | 140 | 140 |
| **Downstream infections averted (assuming average 4 downstream infections per case)** | 1335 | 1530 | 1172 | 1424 | 1330 | 1199 |
| **False positives treated** | 6 | 3 | 450 | 9 | 5 | 300 |

### 4. Additional supplemental results

**Table A4: Variation (range across 10 repetitions) in transmission and clinical benefits of testing at different simulated patient population sizes.** Simulations are run 10 times for the specified population size in the hospital setting. The range (minimum and maximum values) of the output is shown. Population size refers to the number of simulated patients with COVID-19. (Patients who do not have COVID are included in the net benefit calculation, in addition to this number of COVID-19 patients).

| | **Number of COVID-19 patients simulated** | | | |
|---|---|---|---|---|
| | **1,000** | **10,000** | **100,000** | **1,000,000** |
| **Proportion of transmission averted (IQR), RDT** | 0.29 – 0.34 | 0.294 – 0.310 | 0.301 – 0.306 | 0.302 – 0.304 |
| **Proportion of transmission averted (IQR), NAAT** | 0.32 – 0.38 | 0.339 – 0.362 | 0.345 – 0.351 | 0.348 – 0.350 |
| **Proportion of avertible morbidity and mortality averted, RDT** | 0.37 – 0.40 | 0.382 – 0.393 | 0.385 – 0.388 | 0.386 – 0.387 |
| **Proportion of avertible morbidity and mortality averted, NAAT** | 0.42 – 0.44 | 0.423 – 0.434 | 0.425 – 0.428 | 0.426 – 0.427 |

**Figure A3: Potential of diagnostic testing to avert transmission from index cases.** The first row accounts only for assay sensitivity, as measured during acute illness. Then, moving downward, comparisons take into account the timing of presentation; the correlation between detectability and infectivity; and the time from testing to a diagnostic result. (Our final model results also account for empiric isolation before results and incomplete isolation after results, not show here.) **Panel A:** Outpatient setting. Although Ag-RDT only detects 63/80 = 79% of the patients who are NAAT-positive at presentation, those detectable by Ag-RDT account for 77/90 = 86% of future transmission by NAAT-positive patients. Greater delay for NAAT results further increases the relative ability of an Ag-RDT-based approach to avert transmission. **Panel B:** Hospital setting. A longer time from symptom onset to hospital presentation reduces detection by both assays, but the relatively short turnaround time assumed for NAAT (3 days, compared to 1 day in the outpatient setting) compensates for this delay and narrows the gap between Ag-RDT and NAAT.

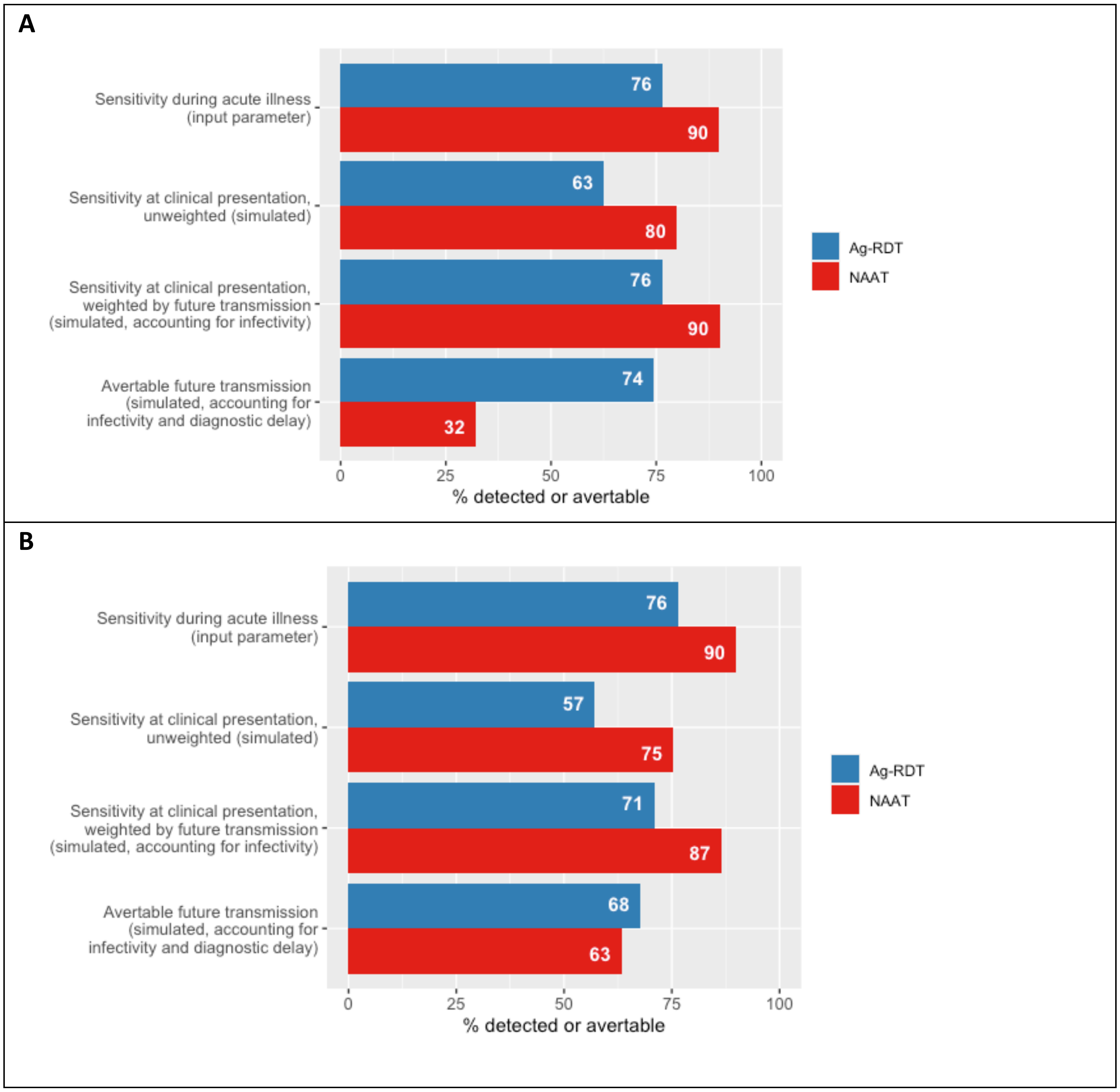

**Figure A4, Decision curve analysis using a standard approach in which a uniform net benefit is accrued for every true-positive case that receives intervention.** Unlike the modified approach shown in Figure 3, these net benefit estimates do not account for (1) the higher infectivity or earlier stage of illness of patients detected by a less sensitive virological assay (Ag-RDT), (2) the benefit lost from diagnostic delay when an assay has a long turnaround time (NAAT), or (3) the potential reduction in intensity of intervention when diagnosis is made with low certainty due to use of a low-specificity test (clinician judgment). Thus, this conventional approach does not detect the advantages of Ag-RDT that cause it to outperform NAAT (and clinician judgment) in the outpatient setting and approach the net benefit of NAAT in the hospital setting.

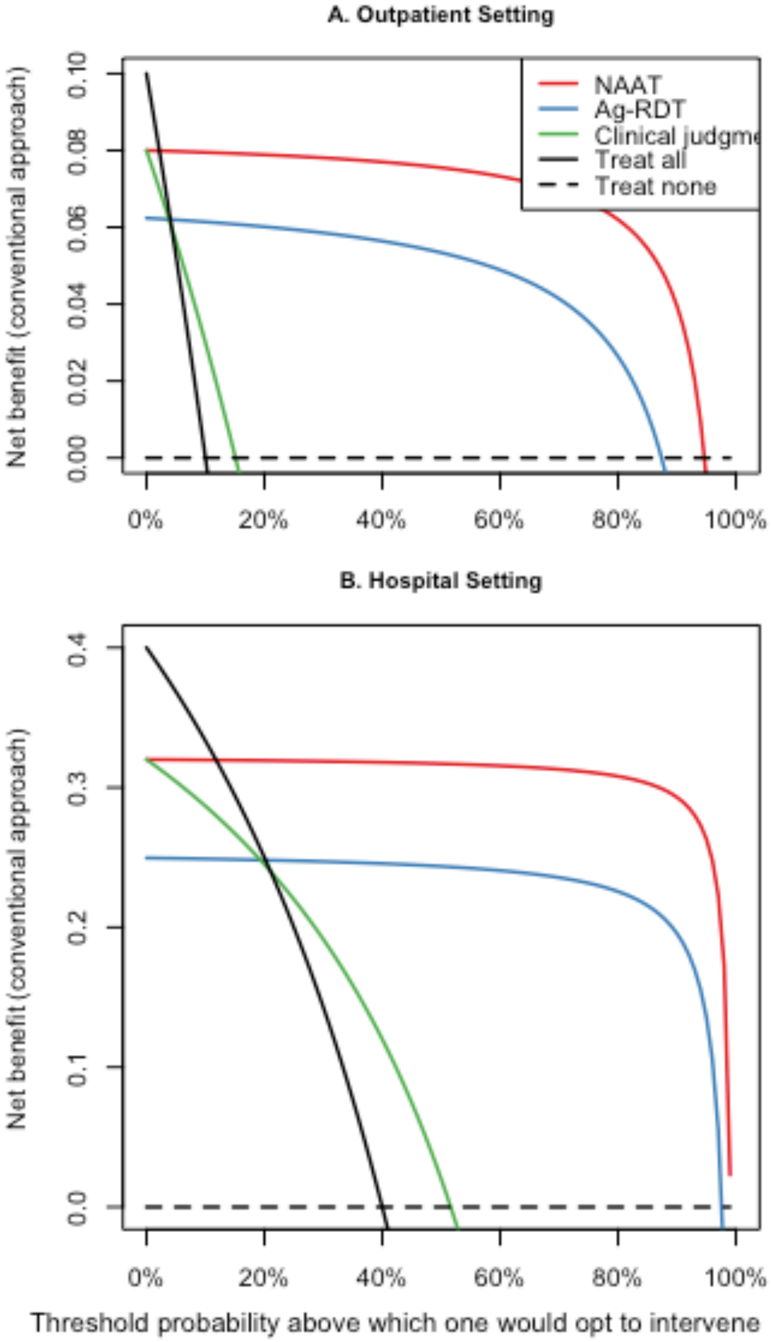
